# The phenotypic changes of γδ T cells in COVID-19 patients

**DOI:** 10.1101/2020.04.05.20046433

**Authors:** Lei Lei, Hongbo Qian, Xiaofang Yang, Xiaobo Zhou, Xingzhe Zhang, Dan Zhang, Tongxin Dai, Rui Guo, Lin Shi, Yanbin Cheng, Baojun Zhang, Jinsong Hu, Yaling Guo

## Abstract

A novel pneumonia-associated respiratory syndrome named coronavirus disease-2019 (COVID-19), which caused by SARS-CoV-2 and broken in Wuhan, China in the end of 2019. Unfortunately, there is no specific antiviral agent or vaccine available to treat SARS-CoV-2 infections. Also, information regarding the immunological characteristics in COVID-19 patients remains limited. Here we collected the blood samples from 18 healthy donors (HD) and 38 COVID-19 patients to analyze changes in γδ T cells. In comparison to HD, the γδ T cells percentage was decreased. γδ T cells are able to immediately respond to SARS-CoV-2 infection and upregulate the activation marker CD25. In addition, the increased expression of CD4 in γδ T cells may serve as a biomarker for the assessment of SARS-CoV-2 infection.

---

A severe pneumonia-associated respiratory syndrome spread rapidly in Wuhan, China in the end of 2019. A novel coronavirus, officially named severe acute respiratory syndrome coronavirus 2, SARS-CoV-2, was identified as the culprit pathogen [1, 2]. It has also been officially named as coronavirus infection disease-19 (COVID-19) by WHO, which listed the outbreak as a public health emergency of international concern. The virus has so far caused 81,896 confirmed cases and 3,287 deaths in China according to WHO. COVID-19 has rapidly spread in more than 180 countries worldwide, including Italia, Iran, Japan, and the United States.

According to the transcriptome sequencing, SARS-CoV-2 shares 74.9% homology with SARS-CoV [3] and 96% homology with a bat coronavirus at the whole-genome level, respectively [2]. According to the pairwise protein sequence analysis of seven conserved non-structural proteins domains, SARS-CoV-2 is an enveloped positive-sense RNA virus, which belongs to the family of coronaviruses including SARS-CoV and MERS-CoV [4, 5].

Currently, there is no specific antiviral agent or vaccine available to treat SARS-CoV-2 infections. Clinical treatments for COVID-19 patients are primarily supportive and symptomatic treatments. Although there are several potential therapeutic targets to repurpose the existing antiviral agents or develop effective interventions against this novel coronavirus [6], toxicology and clinical trials are required for potential uses in the clinic. According to the pathological reports for COVID-19, it was shown that SARS-CoV-2 mainly caused inflammatory responses in the lungs [7]. Several studies showed that COVID-19 patients developed lymphopenia and rising pro-inflammatory cytokines in severe cases [8, 9]. Inflammation can be triggered when innate and adaptive immune cells detect SARS-CoV-2 infection. Innate T cells can provide a first line of defense against pathogens. However, how innate T cells respond to SARS-COV-2 infection remains unclear.

Among innate immune cells, γδ T cells proliferate rapidly and respond to pathogens by inducing apoptosis, mediating antigen presentation and immune regulation [10]. In healthy adult humans, γδ T cells represent 1%-10% of the total circulating lymphocytes, predominately displaying the CD4 and CD8 double negative phenotype [11]. However, in some cases, a fraction of γδT cells can express either CD4 or CD8 [12-14]. γδ T cells do not recognize classical peptide antigens, their TCRs are non-MHC restricted and they can respond to pathogen-associated molecular patterns and produce cytokines in the absence of TCR ligands [15].

In many infections, the number of γδ T cells increases both locally and systemically a few days post-infection. A study found that the ratio of γδ T cells among total lymphocytes in the lungs significantly increased in mice infected with influenza A (H1N1) virus three days post infection [16]. This observation suggests that γδ T cells play an important role in the host immune response. During acute HIV infection, previous studies showed that the surface expression of the activation marker, CD25, is significantly increased on γδ T cells [14], whereas various viruses may have different effects on the patterns of γδ T cells activation [17, 18].

To demonstrate how γδ T cells behave upon SARS-CoV-2 infection, we analyzed the PBMC samples from 38 patients and focused on the characterization of γδ T cell phenotypes. We showed that upon infection, the percentage of γδ T cells in the peripheral blood isolated from COVID-19 patients was drastically decreased when compared with healthy donors (Figure 1A). Although in the acute or early stages of other viral infections, the percentage of γδ T cells increased, we observed a decrease of γδ T cells in symptomatic patients. This might be explained that various types of virus impact γδ T cells in different ways. It is likely that γδ T cell response, including proliferation and cellularity, is dependent on the specific types of viral infections. It is also possible that most patients in this study showed mild symptom besides fever, as opposed to serious illness featuring pneumonia.

**Figure 1.**
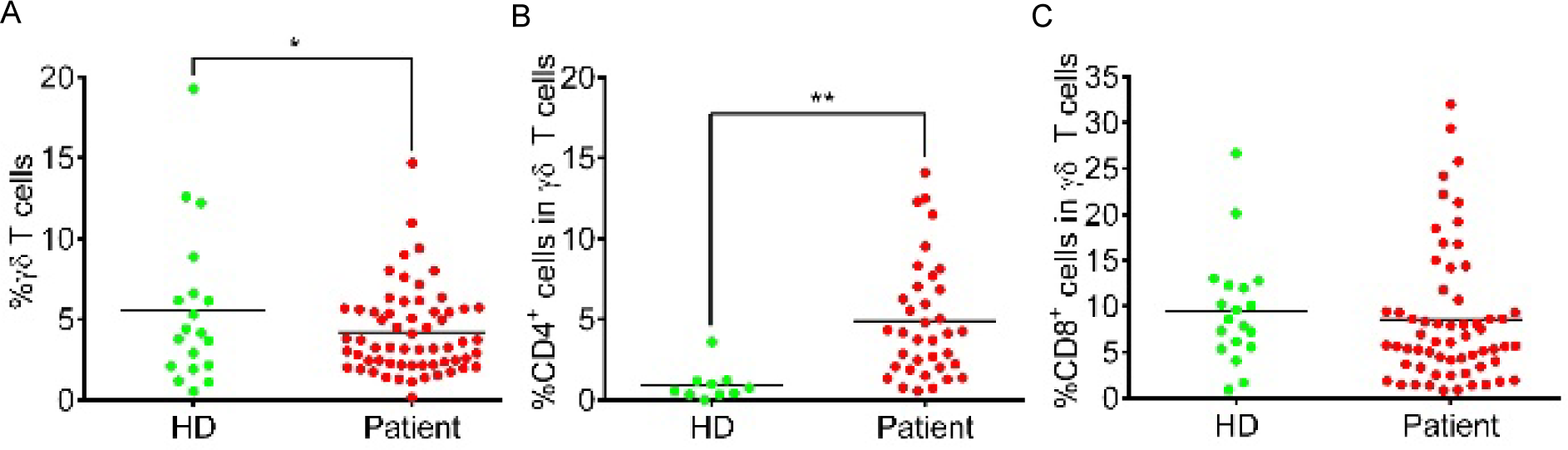
The percentage changes of γδ T cell population in blood of COVID-19 patients and healthy donors. (A) The percentage of γδ T lymphocytes in total blood cells. (B) The percentage of CD4+ γδ T cells in γδ T lymphocyte population. (C) The percentage of CD8+ γδ T cells in γδ T lymphocyte population. Each dot represents a single patient of COVID-19 or healthy donor. * P<0.05 was considered statistically significant.

Interestingly, we found that in comparison to HD group, the percentages of CD4 γδ T cells within the γδ T cell population increased dramatically, while CD8 γδT remained unchanged in COVID-19 patients (Fig1 B and C). The increase of CD4 γδ T cells indicated that in response to SARS-CoV-2 infection, this particular subset of γδ T cells may play roles in antigen presentation and facilitation of activation of adaptive immune cells, which has been demonstrated in different models [19]. The data also suggested this subset of immune cells could immediately respond to viral infection similar to other innate immune cells such as macrophages and dendritic cells, to provide the first line of immune defense. Therefore, γδT cells may act as a bridge between innate and adaptive immunity in response to SARS-CoV-2 infection[20].

In COVID patients, we further observed that γδ T cells exhibited a strong activation phenotype in COVID-19 patients based on CD25 expression (Fig2 B). However, the early activation marker CD69 showed no difference between the patients and healthy donor (HD) group (Fig2 A). It is possible CD69 is expressed strongly earlier during infection, followed by reversion to the quiescent state during prolonged recovery. Since we observed a decreased percentage of γδT cells, we suspected whether γδT cells underwent exhaustion. However, the expression of PD-1 did not differ in γδT cells between HD and COVID-19 patients (Fig2 C).

**Figure 2.**
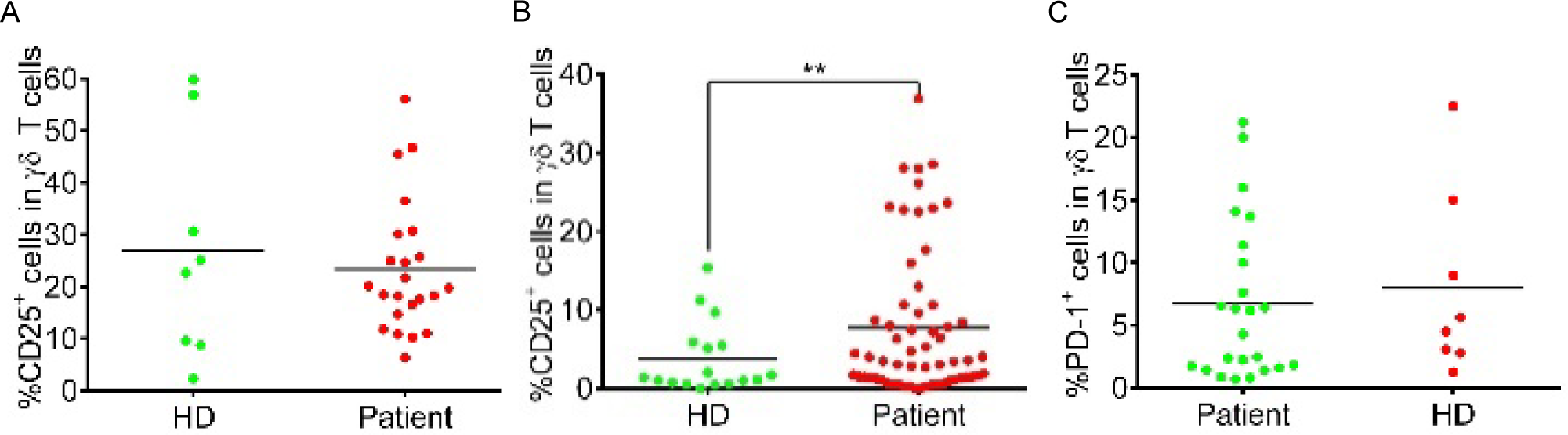
An increase of activated γδ T cells in patients. (A) The percentage of CD69+ cells in γδ T cells. (B) The percentage of CD25+ cells inγδ T cells. (C) The percentage of PD-1+ cells in γδ T cells. Each dot represents a single patient of COVID-19 or healthy donor. * P<0.05 and P<0.01 was considered statistically significant and extremely significant, respectively.

In summary, γδ T cells are able to immediately respond to SARS-CoV-2 infection and upregulate the activation marker CD25. γδ T cells may act in parallel to other innate cells to mediate both direct and indirect defenses against SARS-CoV-2. In addition, the increased expression of CD4 in γδ T cells may serve as a biomarker for the assessment of SARS-CoV-2 infection.

## Materials and Methods

### Ethics statement

This study was approved by the Research Ethics Commission of the Eighth Hospital of Xi’an (20190730-1346). All subjects signed informed consent forms upon admission to hospital. In this study, all cases were taken from the Eighth Hospital of Xi’an (Xi’an, Shaanxi Province, People’s Republic of China).

### Patients

The study included 18 healthy controls and 40 patients from February 18 to March 4. The 38 patients enrolled were all confirmed to have SARS-CoV-2 infection using PCR tests on throat swab specimens.

### Flow cytometry analysis

The Abs used in the flow cytometry analysis are as follows: FITC anti-human TCR γ/δ (B1), APC/Cyanine7 anti-human CD4 (OKT4), PerCP/Cyanine5.5 anti-human CD8 (SK1), APC anti-human CD25 (BC96), PE anti-human CD69 (FN50), APC anti-human CD279 (PD-1) (EH12.2H7) were purchased from Biolegend. Blood cells were stained with Abs in the dark at room temperature for 15 min, and analyzed on a FACSCanto II flow cytometer (BD Biosciences). FlowJo 8 was used for data analysis.

### Statistical analysis

The student’s t test was performed for two group analysis using GraphPad Prism 7.0 software. *\** and *\*\** stands for *P*<0.05 and *P*<0.01, respectively.

## Data Availability

Correspondence and requests for all data should be addressed to Baojun Zhang

## ACKNOWLEDGEMENT

This work was supported in part by grants from Natural Science Foundation of China (NSFC, Grant No. 81820108017), Natural Science Foundation of China (NSFC, Grant No. 81771673), and a COVID-19 special project from Xi’an Jiaotong University Foundation (xzy032020002). We thank all the doctors, nurses, public health workers, and patients for their contribution against SARS-CoV-2 infection.

## Declaration of interests

We declare no competing interests.

## Author contributions

Lei Lei, Xiaofeng Yang, Xingzhe Zhang, Dan Zhang wrote the manuscript. Xiaofang Yang, Tongxin Dai and Rui Guo collected samples and information from patients. Jinsong Hu analyzed FACS data. Xiaobo Zhou, Lin Shi and Yanbin Cheng were responsible for discussing the data. Baojun Zhang generated the idea, designed the experiment, analyzed the data and wrote the manuscript. All authors agree to be accountable for own part of the work.

## Notes

### Competing Interest Statement

The authors have declared no competing interest.

### Funding Statement

This work was supported in part by grants from Natural Science Foundation of China (NSFC, Grant No. 81820108017 and 81771673) and a COVID-19 special project from Xi’an Jiaotong University Foundation (xzy032020002).

